# Multispectral Imaging for MicroChip Electrophoresis Enables Point-of-Care Newborn Hemoglobin Variant Screening

**DOI:** 10.1101/2021.08.05.21261678

**Authors:** Ran An, Yuning Huang, Anne Rocheleau, Alireza Avanaki, Priyaleela Thota, Yuncheng Man, Zoe Sekyonda, Catherine I. Segbefia, Yvonne Dei-Adomakoh, Enoch Mensah, Isaac Odame, Umut A. Gurkan

**Affiliations:** Department of Mechanical and Aerospace Engineering, Case Western Reserve University, Cleveland, OH, USA; HemexHealth, Inc, Portland, OR, USA; Department of Biomedical Engineering, Case Western Reserve University, Cleveland, OH, USA; Department of Child Health, University of Ghana Medical School, Accra, Ghana; Korle Bu Teaching Hospital, Accra, Ghana; Department of Hematology, University of Ghana Medical School, Accra, Ghana; Division of Hematology/Oncology, The Hospital for Sick Children, Toronto, Ontario, Canada; Department of Pediatrics, University of Toronto, Toronto, Ontario, Canada

**Keywords:** Newborn screening, genetic hemoglobin disorders, sickle cell disease, multispectral imaging, point-of-care diagnostics

## Abstract

Hemoglobin (Hb) disorders affect nearly 7% of the world’s population. Globally, around 400,000 babies are born annually with sickle cell disease (SCD), primarily in sub-Saharan Africa where morbidity and mortality rates are high. Although treatments are available for Hb disorders, screening, early diagnosis, and monitoring are not widely accessible due to technical challenges and cost, especially in low-and-middle-income countries. We hypothesized that multispectral imaging will allow sensitive hemoglobin variant identification in existing affordable paper-based Hb electrophoresis, which is a clinical standard test for Hb variant screening. To test this hypothesis, we developed the first integrated point-of-care multispectral Hb variant test: Gazelle-Multispectral. Here, we evaluated the accuracy of Gazelle-Multispectral for Hb variant newborn screening in 321 completed tests in subjects younger than 6 months with known hemoglobin variants including hemoglobin A (Hb A), hemoglobin F (Hb F), hemoglobin S (Hb S) and hemoglobin C (Hb C). Gazelle-multispectral detected levels of Hb A, Hb F, Hb S, and Hb C, demonstrated high correlations with the results reported by laboratory gold standard high performance liquid chromatography (HPLC) at Pearson Correlation Coefficient = 0.97, 0.97, 0.89, and 0.94. Gazelle-multispectral demonstrated 100% sensitivity and 100% specificity in both disease vs normal and disease vs trait, 98.1% sensitivity and 97.0% specificity in trait vs normal in comparison to HPLC in newborns. The ability to obtain rapid and accurate results on newborn samples suggest that Gazelle-Multispectral is suitable for large-scale newborn screening and potentially for accurate diagnosis of SCD in low resource settings.

## INTRODUCTION

Hemoglobin (Hb) disorders are among the world’s most common monogenic diseases. Nearly 7% of the world’s population carry Hb gene variants [1, 2]. Sickle cell disease (SCD) arises when hemoglobin variant mutations are inherited homozygously (HbSS) or paired with another β-globin gene mutation [3, 4]. Manifestations of SCD include acute painful episodes, stroke, chronic hemolytic anemia, organ dysfunction, and early mortality [5]. Globally, an estimated 400,000 babies are born annually with SCD and 70%-75% are in sub-Saharan Africa (SSA) [6-8]. It is estimated that 50-90% of patients with SCD in SSA die by their 5^th^ birthday while 70% of these deaths are preventable [9-11]. Epidemiological modeling shows that universal screening could save the lives of up to 9.8 million newborns with SCD by 2050 with 85% born in SSA [1]. The World Health Organization (WHO) estimates that early diagnosis of SCD coupled with intervention programs would prevent 70% of existing SCD mortality [12].

Effective management of SCD involves genetic counseling, early diagnosis through newborn screening and comprehensive care [13-16]. Newborn screening is the most important public health initiative. SCD newborn screening performed in centralized laboratories has dramatically reduced SCD mortality in resource-rich countries [9, 17]. However, in sub-Saharan Africa and central India, where > 90% of annual SCD births occur, newborn screening programs have not been implemented universally, if at all, due in large part to the cost and logistical burden of laboratory diagnostic tests [18].

SCD newborn screening requires sensitive detection of low levels of certain Hb variants in the context of high levels of expression of other Hb variants. For example, among newborns, normal hemoglobin A (Hb A) and sickle hemoglobin S (Hb S) are expressed at lower levels while fetal hemoglobin (Hb F) is highly expressed making up to 90% of total Hb [19]. The current centralized tests used for newborn screening of SCD are high performance liquid chromatography (HPLC) and isoelectric focusing (IEF)., These tests rely on unaffordable ($15k - $35k) specialized instruments, laboratory facilities, and highly trained personnel, which are lacking in low resource settings where SCD is most prevalent [20]. IEF is a less expensive central test option but lacks quantification capacity, misses certain Hb variants and requires skilled interpretation. Major hospitals in low-resource settings may have access to manual electrophoresis devices but processing samples with these devices are time consuming, need a laboratory setting, require expertise to read, therefore suffers from relatively slow turnaround of test results and start of treatment. Overall, these relatively advanced laboratory techniques require trained personnel and state-of-the-art facilities, which are lacking or in short supply in countries where the prevalence of hemoglobin disorders is the highest [21, 22]. As a result, there is a need for affordable, portable, easy-to-use, accurate, point-of-care (POC) tests to facilitate decentralized hemoglobin testing in low-resource settings to enable nationwide newborn screening.

Several POC diagnostic systems for SCD have been described based on testing methods such as sickle cell solubility test and antibody-based lateral flow assays such as Sickle SCAN™ and HemoType SC™ [23-25]. However, the sickle cell solubility test is not reliable for samples with Hb S levels below 20% [23]. As a result, this method is not suited for screening for Hb S in newborns, where Hb S levels are normally below 20% [32, 33][26]. Antibody-based lateral flow assays only report qualitative instead of quantitative test results [24]. Additionally, most of these tests only detect three hemoglobin variants (Hb A, Hb S, and Hb C) while missing Hb F and others, which may result in compromised detection sensitivity and specificity. Moreover, these tests lack portable readers for objective analysis or electronic record keeping, and rely on visual interpretation and manual recording of the test results, which is prone to significant errors in the field [27]. In fact, user misinterpretation and data entry errors have been reported to range between 2.3% to 26.9%, which can significantly compromise the accuracy of these tests [25].

In a 2019 report, the World Health Organization (WHO) listed hemoglobin testing as one of the most essential in vitro diagnostic (IVD) tests for primary care use in low and middle income countries [28]. Furthermore, hemoglobin electrophoresis has recently been added to the WHO essential list of IVDs for diagnosing SCD and sickle cell trait [2]. Leveraging the WHO recognized Hb electrophoresis test, we developed a paper-based, miniaturized Hb electrophoresis platform, Gazelle™ [3, 4, 21] (**Fig.1**).

Gazelle™ has been extensively tested and validated for hemoglobin variants, including SCD and hemoglobin E disease in the US, India, and several countries in Africa and Southeast Asia [4][22].

Here, we implemented multispectral imaging under both white illumination and ultraviolet (UV) illumination at 410 nm wavelength and developed the Gazelle-Multispectral platform as the first POC test able to identify and quantify Hb variants in newborns and people of any age. The high absorbance of hemoglobin at 410 nm wavelength enhances the limit of detection thus allows detection and identification of hemoglobin variants at low concentrations, which is crucial for SCD screening in newborns. In this manuscript, we firstly determined the limit of detection of the Gazelle-Multispectral platform in laboratory environment using controlled samples in Portland, OR, USA. Additionally, we describe a study for evaluating the diagnostic performance of this platform for screening HbSS, HbSC disease, and the related carrier states (Hb S trait and Hb C trait) using whole blood at the POC in 321 completed tests in Korle Bu, Ghana, a location selected for its high prevalence of both the Hb S and Hb C variants. The limit of detection of Gazelle-Multispectral was determined to be 4%. This low limit of detection is sufficient for detecting the low percentage of Hb S typically found in newborn samples for newborns with sickle cell trait and SCD. The clinical test results demonstrate Gazelle-Multispectral determines both disease vs normal and disease vs trait at 100% sensitivity and 100% specificity, as well as determines trait vs normal at 98.1% sensitivity and 97.0% specificity. Overall, the ability to achieve low limit of detection and the ability detect Hb variant accurately and rapidly in newborns suggest that Gazelle-Multispectral is suitable for large-scale newborn screening and potentially for accurate diagnosis.

## METHODS

### Laboratory Determination of Limit of Detection

Gazelle Multispectral’s lower limit of detection for Hb S was determined using artificially created samples by mixing a heathy donor’s blood sample with known Hb A level (96.3% Hb A, 0.4% Hb F, 3.3% Hb A_2_, determined by HPLC) and a blood sample from a patient with SCD undergoing transfusion therapy with known levels of Hb A and Hb S (59.1% Hb A, 39.3% Hb S, 0.2% Hb F, 1.4% Hb A_2_, determined by HPLC). Hb S levels in the artificially created samples were obtained by HPLC as 39.3%, 19.2%, 12.8%, 9.6%, 6.4%, 3.2%, 1.6%, and 0.8%. Each artificially created sample was tested 3 times using Gazelle-Multispectral and the reported results were compared with Hb S levels determined by HPLC.

### Study Design and Oversight

We conducted a prospective diagnostic accuracy study on Gazelle-Multispectral for detecting Hb variants including Hb A, Hb F, Hb S, and Hb C at Korle Bu Teaching Hospital (KBTH), Accra, Ghana using Institutional Review Board (IRB) approved protocol (KBTH-IRB/00078/2019). The results obtained using the investigational assay, Gazelle-Multispectral, were compared to the results reported by the reference (“Gold-standard”) tests using HPLC. The Gazelle platform was designed and provided by Hemex Health, headquartered in Portland, Oregon, USA. The laboratory standard test used in the study was HPLC at KBTH. All authors have reviewed and analyzed the data and attest to their accuracy and completeness as well the fidelity of adherence to the study protocol.

### Study Populations and Procedures

This test was conducted at KBTH, the largest public hospital in Ghana. Newborns were enrolled from the postnatal wards, and children were enrolled from the Child Welfare Clinic, during routine immunization visits. The study was approved by the KBTH IRB and informed consent obtained from each participant’s parent or guardian. A blood sample was obtained from each participant using finger prick at the vaccination clinic or heel prick for newborns. Any blood samples not tested immediately on Gazelle-Multispectral were refrigerated until use. After a Gazelle test was conducted, the remaining blood from the blood collection tube was saved and frozen at -80 °C. HPLC tests were performed at KBTH on the frozen samples using the D-10 HPLC system (Bio-Rad Laboratories, Hercules, CA, USA).

### Gazelle Multispectral Test Procedure

The technician performed the tests according to the Gazelle-Multispectral instructions for use as published previously [4]. Briefly, 20 µl of blood and 40 µl of Gazelle Marker Fluid was pipetted into an Eppendorf tube which was then vortexed for 20 seconds. 50 µl of Gazelle Buffer was used to wet the Gazelle Hb Variant Cartridge paper and the cartridge was soaked for 1 minute. 20 µl of the blood mixture was pipetted onto a glass slide and a customized stamper was touched to the mixture. The blood sample was wicked and filled the stamper completely. The stamper stand was placed directly over the cartridge and the stamper with blood and marker was placed into the stamper stand. The operator held down the stamper stand for 5 seconds to apply the blood and marker mixture to the cartridge. The cartridge was flipped over and 200 µl were pipetted into the wells on each end of the cartridge. Finally, the cartridge was placed into the Gazelle Reader and the test was started. After 8 minutes, the results screen showed the percentages of each hemoglobin type present in the blood sample as well as an interpretative statement.

### Confirmatory Laboratory Procedures

Blood samples that were stored at -80°C were retrieved and thawed. 5 µl of sample was pipetted and diluted with 1500 µl of distilled water. Diluted hemolysates were arranged on racks and loaded into the Bio-Rad D-10 HPLC system. Each sample was tested for approximately 6 minutes. The results reported for each blood sample included the relative percentages of each hemoglobin type present.

### Gazelle-Multispectral Data Analysis

Customized data analysis algorithm was integrated in Gazelle-Multispectral system. This data analysis algorithm automatically identifies sickle cell disease (FS, FSC, FSA), sickle cell trait (FAS), Hemoglobin C Trait (FAC), and normal phenotype (FA) based on Hb band migration pattern as described previously [4]. The data analysis algorithm also automatically quantifies the relative percentages of Hb A, Hb F, Hb S, and combined Hb C. The Gazelle-Multispectral reported Hb variant identification and quantification results were compared with the ones reported by HPLC using Pearson correlation and Bland-Altman analysis. Gazelle-Multispectral sensitivity, specificity, positive predictive value (PPV, and negative predictive value (NPV) in identification of SCD (FS/FSC/FSA) vs. Normal (FA), SCD (FS/FSC/FSA) vs.

Sickle Cell Trait (FAS) and Hemoglobin C Trait (FAC), and Sickle Cell Trait (FAS) and Hemoglobin C Trait (FAC) vs. Normal (FA) were calculated for the study population compare to HPLC reported results.

The primary objective was to determine the limit of detection for detecting individual Hb variants, as well as the sensitivity, specificity, PPV, and NPV, of Gazelle Multispectral, compared to reference tests, in detecting normal Hb (Hb A), fetal Hb (Hb F), and common pathologic Hb variants (Hb S and Hb C), in whole blood specimens from newborns and older children. The main goal was to test the ability of Gazelle Multispectral to accurately detect HbSS, HbSC disease, and the related carrier states (Hb S trait and Hb C trait) in newborns and older children.

## RESULTS

### Test Population

A total of 321 completed tests from both Gazelle-Multispectral and HPLC acquired at KBTH, Ghana were included in this study. We defined ‘Valid’ and ‘Inconclusive’ tests according to published recommendations in literature [29] and the STARD guidelines [30]. A ‘Valid’ test was defined as a test that performed as expected according to objective standards and the test result was reported properly from the data analysis algorithm. An ‘Inconclusive’ test was a test that performed adequately according to an objective set of standards. However, an ‘Inconclusive’ test has quantification confidentiality value automatically evaluated by the algorithm that is lower than the preset threshold value, which can be recognized at the end of the test. Reasons for ‘Inconclusive’ tests include appearance of a band or bands at or close to the borderline region between two adjacent detection windows or with results that were not consistent with any of the standard Gazelle interpretive statements. In this study, 294 out of 321 (91.6%) tests were recognized as ‘Valid’, while 27 out of 321 (8.4%) tests were recognized as ‘Inconclusive’.

### Gazelle-Multispectral separates, images, and tracks hemoglobin variants real-time under multi-spectrum during electrophoresis

The fundamental principle behind Gazelle-Multispectral technology is hemoglobin electrophoresis, in which different (bio)molecules including total hemoglobin, standard calibrator, and hemoglobin variants can be separated based on their charge-to-mass ratio when exposed to an electric field in the presence of a carrier substrate. Gazelle-Multispectral is single-use and cartridge-based, which can be mass-produced at low-cost (**Fig. 1A**) [4]. Tris/Borate/EDTA (TBE) buffer is used to provide the necessary ions for electrical conductivity at pH of 8.4 in the cellulose acetate paper. Hb molecules carry net negative charges under this pH and caused them to travel from the cathode to the anode upon exposure to electric field. The electric mobility differences of various hemoglobin phenotypes allow separation and thus identification of each hemoglobin variant. Separated hemoglobin variants are imaged under both white light illumination (**Fig. 1B**) and ultraviolet (UV) light illumination (**Fig. 1C**). The acquired data under white light illumination demonstrates the natural red color of hemoglobin and thus validates the Gazelle-Multispectral tests (**Fig. 1B)**. The acquired data under UV light illumination is used for sensitive and accurate identification and quantification of Hb variants (**Fig. 1D&E**). Data acquired under UV illumination yields an enhanced limit of detection and a higher signal to background ratio than white light illumination data. Combining both white light and UV detection spectrums, Gazelle-Multispectral automatically tracks, detects, identifies and quantifies electrophoretically separated low concentration hemoglobin variants.

**Figure 1.**
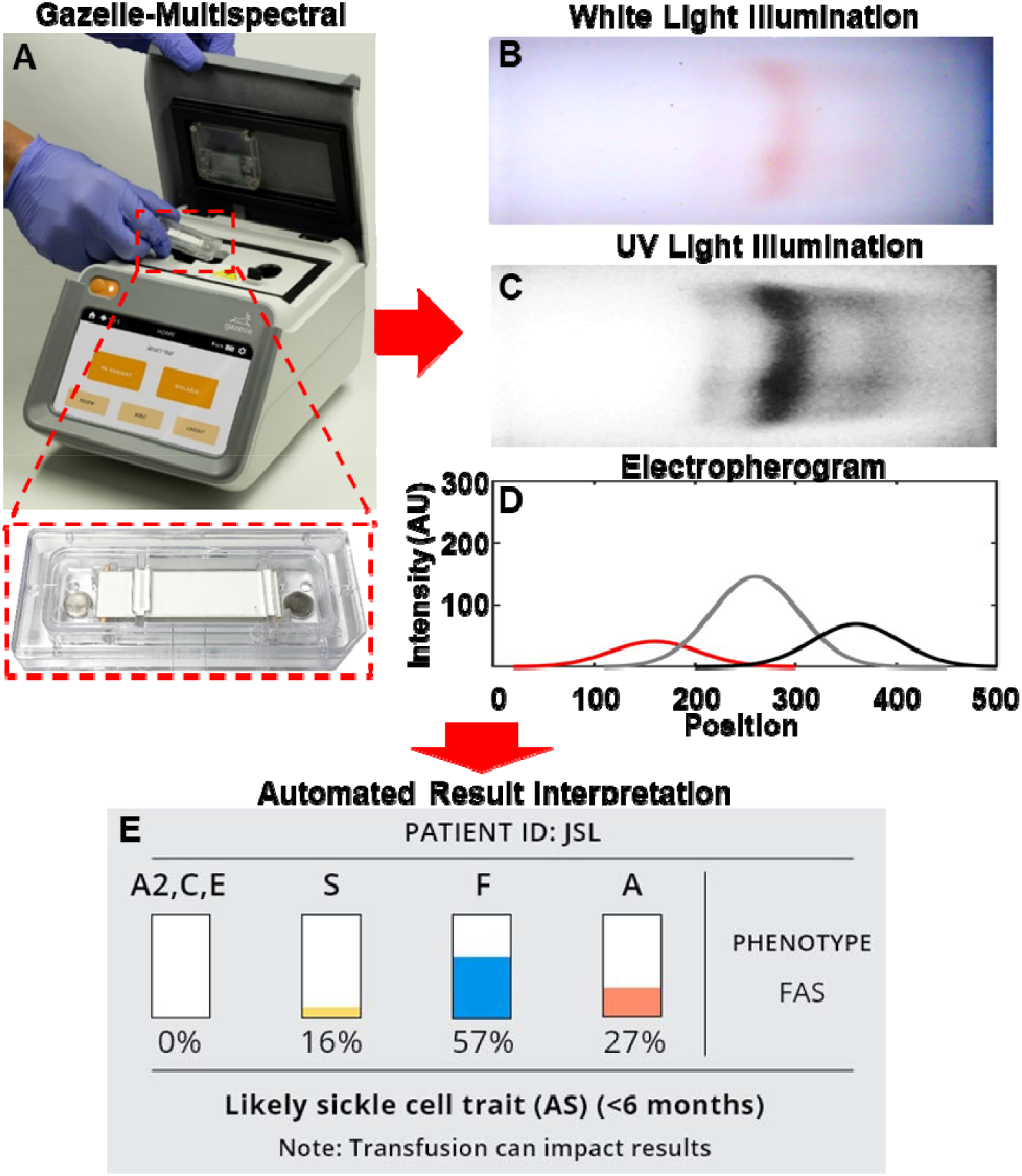
Gazelle-Multispectral for screening hemoglobin variants in newborns. **(A)** Gazelle-Multispectral platform for paper-based microchip electrophoresis using disposable cartridge **(red box)** at the point of need. The Gazelle-Multispectral perform real time imaging and data analysis tracking the Hb electrophoresis process under both white light illumination **(B)** and UV light illumination **(C)**. The images captured under white light illumination provides visual validation of test progression. **(D)** The images captures under UV illumination are used for identifying and quantifying Hb variants are in real time using an internally integrated data analysis algorithm. **(E)** At the end of each test, Gazelle-Multispectral algorithm automatically reports the identification and quantification of Hb variant results, and determines the patient phenotype accordingly.

### Analysis of limit of detection

In the analytical assessment, the limit of detection (LOD) was determined as the lowest concentration for which all three replicates scored positive. Gazelle-Multispectral consistently identified Hb S in 3 out of 3 replicates for artificially created samples with Hb S levels at 3.2%, 6.4%, 9.6%, 12.8%, 19.2%, and 39.3%. However, Gazelle-Multispectral identified Hb S in 0 out of 3 replicates for artificially created samples with Hb S levels at 1.6% and 0.8%. As a result, the lower LOD of Gazelle-Multispectral for identifying Hb S was set at 4%. This 4% lower LOD is sufficient for detecting low percentage of Hb S typically found in newborn samples for newborns with sickle cell trait (FAS: Hb S 6.5 ± 2.8%) and SCD (FS: Hb S 10.2 ± 3.9%) [31].

### Gazelle-Multispectral automatically validates electrophoresis separation, identifies low concentration hemoglobin, and determines their relative percentages based on multi-spectrum imaging

Gazelle-Multispectral image tracking and data analysis method is similar to its previous generation, Gazelle, as described in our previous publication [4]. Briefly, Gazelle-Multispectral algorithm recognizes the initial application point of the mixture containing a blue control marker and hemoglobin. The algorithm then distinguishes and tracks the blue marker and hemoglobin according to their naturally distinct blue and red colors within the white light field during separation. The tracked blue marker and red hemoglobin migration pattern is analyzed by the image processing and decision algorithm to determine test validity (**Fig. 2A-H**). A validated test will be further analyzed utilizing data acquired under UV field (**Fig. 2I-T**). A space-time plot was generated to illustrate the entire band migration on the paper (x-axis, from left to right) within the entire time (y-axis, from top to bottom, **Fig. 2I-L**). Four representative tests with different Hb variants were demonstrated in **Fig. 2** (**Column 1**: Healthy newborn, FA; **Column 2**: Newborn with SCD, FS; **Column 3**: Newborn with Sickle Cell Trait, FAS; and **Column 4**: Newborn Hemoglobin C Trait, FAC). Gazelle-Multispectral sensitively and accurately identified Hb variants agreeing with HPLC reported results in all these 4 representative samples (Gazelle-Multispectral vs. HPLC; FA: Hb F: 92% vs. 94%; Hb A: 8% vs. 6%; FS: Hb F 83% vs. 89%, Hb S 17% vs. 11%; FAS: Hb F 57% vs. 55%, Hb A 27% vs. 29%, Hb S 16% vs. 16%; and FAC: Hb F 45% vs. 45%, Hb A 35% vs. 35%, Hb C 20% vs. 20%).

**Figure 2.**
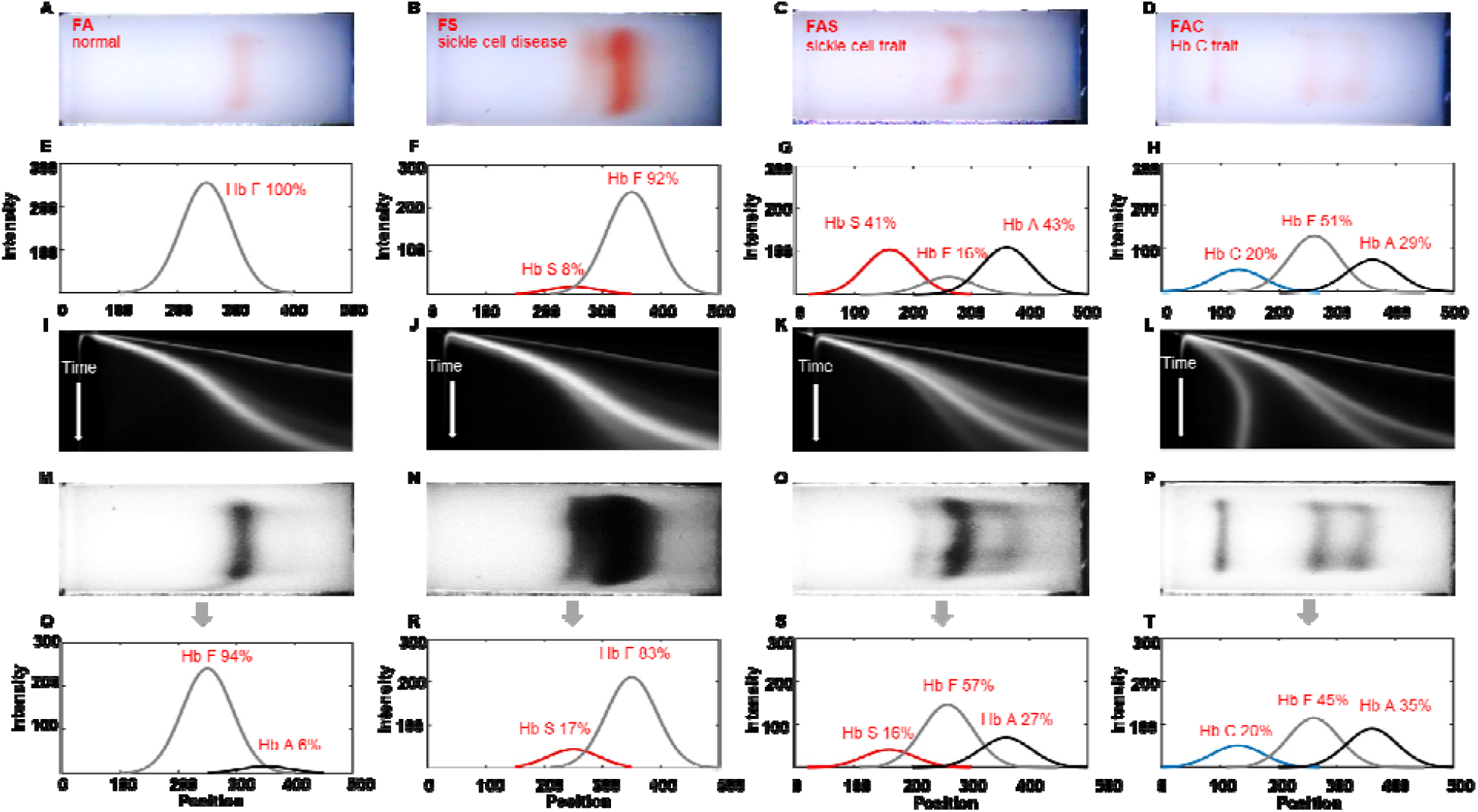
Identification of Hb variants and quantification of Hb percentages by Gazelle-Multispectral. **(A-D)** The first row shows images captured under white light field. **(E-H)** The second row shows electropherograms generated based on the white light images using the white data analysi algorithm. **(I-L)** The third row illustrates 2D representation of Gazelle-Multispectral space-time plots of band migration in UV imaging mode, which are used with the machine learning algorithm. (M-P) The fourth row shows images captured under UV light field. **(Q-T)** The fifth row shows electropherogram generated based on the UV light images using the UV data analysis algorithm. The Gazelle-Multispectral UV data analysis algorithm sensitively and accurately identified Hb variants agreeing with HPLC reported results. Column 1 – 4: Multispectral test results for samples with different phenotypes. Column 1: Hb FA (Healthy newborn, Hb A: 8% vs. 6%, Hb F: 92% vs. 94%, Gazelle-Multispectral vs. HPLC); Column 2: Hb FS (Newborn with sickle cell disease, Hb F 83% vs. 89%, Hb S 17% vs. 11%, Gazelle-Multispectral vs. HPLC); Column 3: Hb FAS (Newborn with sickle cell trait, Hb S 16% vs. 16%, Hb F 57% vs. 55%, Hb A 27% vs. 29%, Gazelle-Multispectral vs. HPLC); and Column 4: Hb FAC (Newborn with Hb C disease, Hb C 20% vs. 20%, Hb F 45% vs. 45%, Hb A 35% vs. 35%, Gazelle-Multispectral vs. HPLC). Gazelle-Multispectral enabled identification and quantification of low concentration Hb variants with higher sensitivity **(I–T)** compared to white light imaging mode **(A-H)**.

### Gazelle-Multispectral Hb variant quantification demonstrated high correlation with HPLC

Pearson correlation analysis and Bland-Altman analysis were performed on the 294 tests recognized as ‘Valid’. The correlation plots include the Gazelle-Multispectral determined Hb variant levels (y axis) versus the Hb variant levels reported by the HPLC (x axis) including Hb A, Hb F, Hb S, and Hb C (**Fig. 3 First Column**). The Bland-Altman analysis plots demonstrate the difference between Gazelle-Multispectral determined Hb variant levels (y axis) at the entire range of Hb levels detected (x axis, **Fig. 3 Second Column**). The results from Pearson correlation analysis demonstrate Pearson correlation coefficient (PCC) of 0.97, 0.97, 0.89, and 0.94 for Hb A, Hb F, Hb S, and Hb C. Bland-Altman analysis showed Gazelle-Multispectral determines blood Hb variant levels with mean bias of 2.3% for Hb A (Limits of agreement, LOA: -9.1% to 13.7%); -2.7% for Hb F (LOA: -16.0% to 10.5%); 0.8% for Hb S (LOA: -5.5% to 7.1%), and -0.3% for Hb C (LOA: -3.0% to 2.3%, **Fig. 3, Second Column**). Together, these results revealed strong agreement between Gazelle-Multispectral determined Hb variant levels and HPLC reported Hb variant levels (**Fig. 3, First Column**).

**Figure 3.**
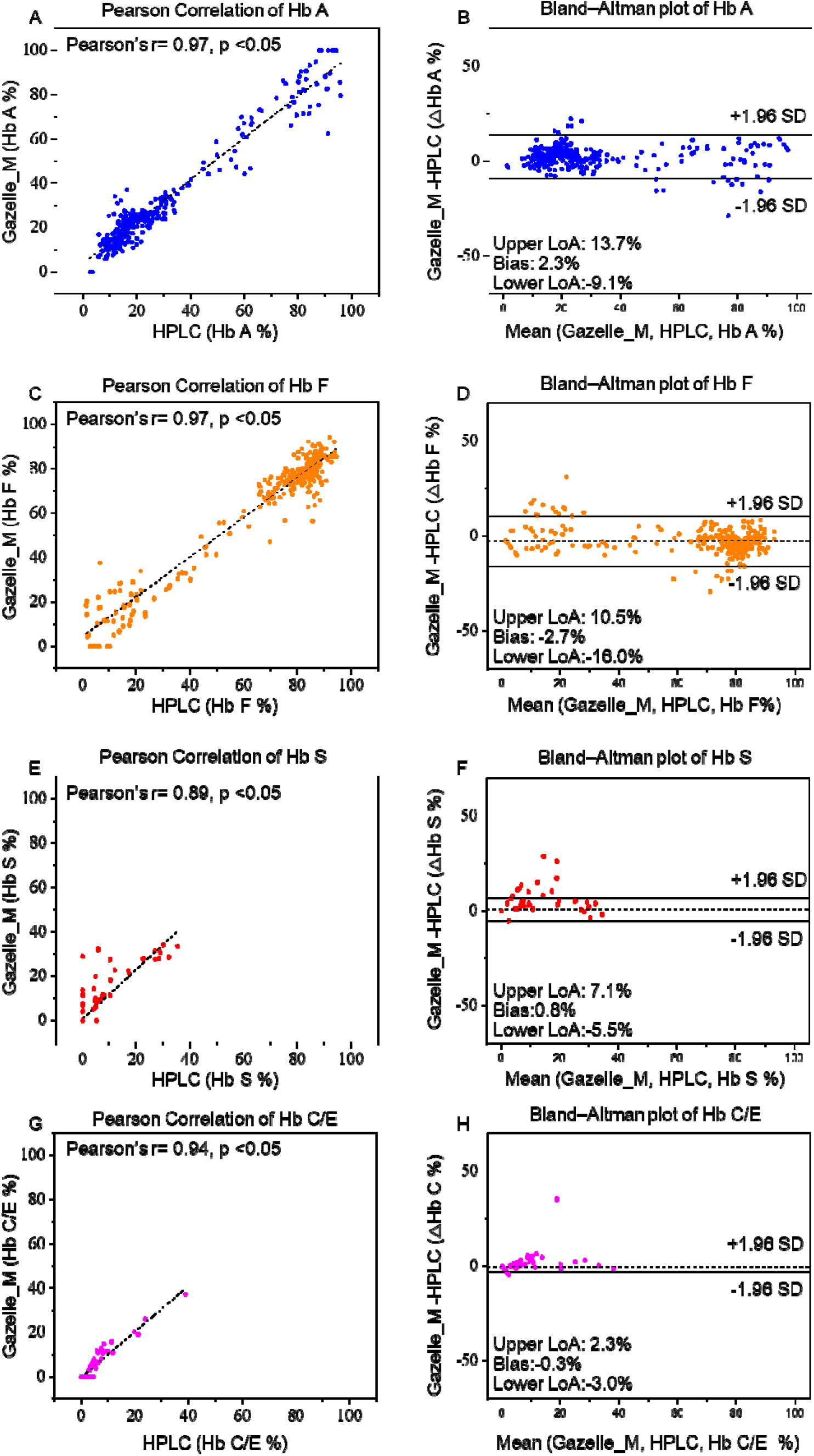
Gazelle-Multispectral Hb variant identification and quantification in 294 newborns. Pearson correlation (Column 1) and Bland-Altman analysis (Column 2) showed Gazelle-Multispectral identified and quantified Hb A (**A&B**, Pearson coefficient correlation (PCC) = 0.97, p < 0.05, Mean bias ± 1.96 × Standard Deviation (SD) = 2.3% ± 11.4%), Hb F (**C&D**, PCC = 0.97, p < 0.05, Mean bias ± 1.96SD = -2.7% ± -13.3%), Hb S (**E&F**, PCC = 0.89, p < 0.05, Mean bias ± 1.96 SD = 0.8% ± 6.2%), and Hb C levels (**G&H**, PCC = 0.94, p < 0.05, Mean bias ± 1.96SD = -0.3% ± 2.7%) agree with the ones reported by laboratory standard HPLC. In Column 2, the solid black lines indicate the mean biases and the dashed gray lines represent 95% limits of agreement.

### Gazelle-Multispectral performs SCD newborn screening at high sensitivity and specificity

In this clinical study, Gazelle-Multispectral test results included the following (**Table 1**): disease (FS/FSC/FSA), Hb S (FAS), Hb C Trait (FAC), and normal (FA). Gazelle-Multispectral identified subjects with disease including FS, FSC, and FSA from normal subjects and subjects with the related carrier states (Hb S trait, FAS and Hb C trait, FAC) with 100% sensitivity, specificity, PPV and NPV (**Table 1**). Seven subjects with normal Hb (FA) were identified as Sickle Cell Trait (FAS Table 1). One subject with Sickle Cell Trait (FAS) was identified as normal Hb (FA, Table 1). Sensitivity, specificity, PPV, and NPV for identifying subjects with the related carrier states from normal subjects (Trait vs. Normal) were 98.1%, 97.0%, 88.3%, and 99.6%.

**Table 1:** Gazelle-Multispectral screening sensitivity, specificity, positive predictive value (PPV), and negative predictive value (NPV) in comparison to reference standard method.

|  | Disease vs. Normal<sup>a</sup> | Disease vs. Trait<sup>b</sup> | Trait vs. Normal<sup>c</sup> |
| --- | --- | --- | --- |
| **True positive, TP** | 3 | 3 | 53 |
| **True negative, TN** | 230 | 53 | 230 |
| **False Positive, FP** | 0 | 0 | 7 |
| **False negative, FN** | 0 | 0 | 1 |
| **Sensitivity, TP/(TP + FN)** | 100.0% | 100.0% | 98.1% |
| **Specificity, TN/(TN + FP)** | 100.0% | 100.0% | 97.0% |
| **PPV, TP/(TP + FP)** | 100.0% | 100.0% | 88.3% |
| **NPV, TN/(TN + FN)** | 100.0% | 100.0% | 99.6% |

<sup>a</sup>SS/SC/FS/FSC vs. AA/FA
<sup>b</sup>SS/SC/FS/FSC vs. AS/AC/FAS/FAC
<sup>c</sup>AS/AC/FAS/FAC vs. AA/FA

## Discussion

Hb F expression represents up to 90% of total Hb in newborns [32, 33]. Gazelle-Multispectral implements multispectral imaging into a point-of-care hemoglobin electrophoresis test, which allows sensitive detection and quantification of low concentration Hb variants with a limit of detection at 4%, thus enables SCD screening for Hb variants among newborns having low levels of Hb A and Hb S. The Gazelle-Multispectral algorithm automatically provides Hb variant identification and quantification results of relative Hb percentages (**Fig. 1E**) and does not require users to perform result interpretation. Additionally, Gazelle-Multispectral uses a low-cost disposable cartridge, does not require a dedicated lab environment, and can be operated on battery power. These critical features distinguish Gazelle-Multispectral from current available laboratory methods and other emerging POC technologies and enable affordable, simple, and rapid newborn screening for Hb variant at the point of care.

In this clinical study conducted among 321 newborns in Korle Bu, Ghana, Gazelle-Multispectral demonstrated 100% sensitivity and specificity for identifying subjects with diseases vs. healthy subjects; and subjects with disease vs. subjects with sickle cell trait and Hb C trait; Additionally, Gazelle-Multispectral demonstrated 98.1% sensitivity and 97.0% specificity for identifying subjects with sickle cell trait and Hb C trait vs. healthy subjects. Common practice in Hb testing is that all positive test results are confirmed with a secondary method prior to final diagnostic decision making and treatment initiation [34]. Therefore, all disease positive tests would likely result in a secondary confirmatory test that should eliminate the small number of false positives.

Hb electrophoresis techniques overall share a common limitation in discriminating certain Hb variants due to their similar electrophoretic mobilities at given condition. For example, it is challenging to discern Hb C and Hb A_2_, even using a laboratory platform of capillary zone electrophoresis because these hemoglobin variants demonstrate partially overlapped peak within the same detection window [35, 36]. This peak overlapping is also a challenge for the reference standard HPLC as well as its alternatives [35, 37, 38]. Notably, Hb C and Hb E are known to co-migrate in paper-based hemoglobin electrophoresis. Therefore, Gazelle-Multispectral reports Hb C/E instead of reporting Hb C or E individually. However, these Hb variants display distinct geographical prevalence and distribution. For example, Hb C is highly prevalent in West Africa thus is related to this study [39], while Hb E is the most prevalent in the Mediterranean region, Southeast Asia, and in the Indian subcontinent [40-44]. As a result, test location, the ethnicity of the subject, and clinical history can be used to facilitate differentiate between these co-migrating Hb types. For example, in this particular study, and all Hb variant recognized as Hb C/E were identified as Hb C due to the test location.

In summary, Gazelle-Multispectral enables affordable and rapid identification of common Hb variants in newborns at the point-of-need. The Gazelle-Multispectral reader provide animated on-screen instructions of step-by-step guidance for test operation procedures to minimize user errors. The internally integrated data analysis algorithm automatically reports Hb variant identification and quantification results in an objective and easily understandable manner. Gazelle-Multispectral is a versatile, mass-producible, multispectral detection-based electrophoresis platform technology for affordable, rapid, and accurate diagnostic testing and newborn screening programs for SCD at the POC in low resource regions where the prevalence of SCD is high.

## Data Availability

All reasonable requests for materials and data will be fulfilled by the corresponding author of this publication.

## ACKNOWLEDGEMENTS

Authors acknowledge National Heart Lung and Blood Institute Small Business Innovation Research Program (R44HL140739, R41HL151015), National Heart Lung and Blood and the National Center for Complementary & Integrative Health (NCCIH) 1U54HL143541, National Institute of Diabetes and Digestive and Kidney Diseases Small Business Innovation Research Program (R41DK119048), and NHLBI R01HL133574 and T32HL134622. This article’s contents are solely the responsibility of the authors and do not necessarily represent the official views of the National Institutes of Health.

## AUTHOR CONTRIBUTIONS

RA, YH, and AA contributed to the proof-of-concept experiments and initial development. AR, PT, CS, YDA, EM, and IO helped with the planning and execution of clinical testing, including human subject research protocol development, subject recruitment, blood sample collection, and testing. RA, YH, AR, AA, YM, and ZS performed the data analysis, prepared the tables, figures, figure captions, and supplementary information. RA drafted the manuscript and all authors edited the manuscript. RA, YH, AR, AA, PT, YM, ZS, CS, YDA, EM, IO, and UAG reviewed and edited the manuscript.

## References

1. Piel, F.B., A.P. Patil, R.E. Howes, O.A. Nyangiri, P.W. Gething, M. Dewi, W.H. Temperley, T.N. Williams, D.J. Weatherall, and S.I. Hay, Global epidemiology of sickle haemoglobin in neonates: a contemporary geostatistical model-based map and population estimates. The Lancet, 2013. 381(9861): p. 142–151.

2. (WHO), W.H.O., Second WHO Model List of Essential In Vitro Diagnostics. 2019(WHO/MVP/EMP/2019.05).

3. An, R., M.N. Hasan, Y. Man, and U.A. Gurkan. Integrated Point-of-Care Device for Anemia Detection and Hemoglobin Variant Identification. in 2019 IEEE Healthcare Innovations and Point of Care Technologies, (HI-POCT). 2019.

4. Hasan, M.N., A. Fraiwan, R. An, Y. Alapan, R. Ung, A. Akkus, J.Z. Xu, A.J. Rezac, N.J. Kocmich, M.S. Creary, T. Oginni, G.M. Olanipekun, F. Hassan-Hanga, B.W. Jibir, S. Gambo, A.K. Verma, P.K. Bharti, S. Riolueang, T. Ngimhung, T. Suksangpleng, P. Thota, G. Werner, R. Shanmugam, A. Das, V. Viprakasit, C.M. Piccone, J.A. Little, S.K. Obaro, and U.A. Gurkan, Paper-based microchip electrophoresis for point-of-care hemoglobin testing. Analyst, 2020. 145(7): p. 2525–2542.

5. Kato, G.J., F.B. Piel, C.D. Reid, M.H. Gaston, K. Ohene-Frempong, L. Krishnamurti, W.R. Smith, J.A. Panepinto, D.J. Weatherall, F.F. Costa, and E.P. Vichinsky, Sickle cell disease. Nature Reviews Disease Primers, 2018. 4: p. 18010.

6. Alapan, Y., A. Fraiwan, E. Kucukal, M.N. Hasan, R. Ung, M. Kim, I. Odame, J.A. Little, and U.A. Gurkan, Emerging point-of-care technologies for sickle cell disease screening and monitoring. Expert Rev Med Devices, 2016. 13(12): p. 1073–1093.

7. McGann, P.T. and C. Hoppe, The pressing need for point-of-care diagnostics for sickle cell disease: A review of current and future technologies. Blood cells, molecules & diseases, 2017. 67: p. 104–113.

8. Hsu, L., O.E. Nnodu, B.J. Brown, F. Tluway, S. King, L.G. Dogara, C. Patil, S.S. Shevkoplyas, G. Lettre, R.S. Cooper, V.R. Gordeuk, and B.O. Tayo, White Paper: Pathways to Progress in Newborn Screening for Sickle Cell Disease in Sub-Saharan Africa. J Trop Dis Public Health, 2018. 6(2): p. 260.

9. El-Haj, N. and C.C. Hoppe, Newborn Screening for SCD in the USA and Canada. International Journal of Neonatal Screening, 2018. 4(4): p. 36.

10. Sabarense, A.P., G.O. Lima, L.M. Silva, and M.B. Viana, Characterization of mortality in children with sickle cell disease diagnosed through the Newborn Screening Program. J Pediatr (Rio J), 2015. 91(3): p. 242–7.

11. Wang, Y., G. Liu, M. Caggana, J. Kennedy, R. Zimmerman, S.O. Oyeku, E.M. Werner, A.M. Grant, N.S. Green, and S.D. Grosse, Mortality of New York children with sickle cell disease identified through newborn screening. Genet Med, 2015. 17(6): p. 452–9.

12. Regional Committee for, A., Sickle-Cell Disease: a strategy for the WHO African Region. 2011.

13. Quarmyne, M.O., W. Dong, R. Theodore, S. Anand, V. Barry, O. Adisa, I.D. Buchanan, J. Bost, R.C. Brown, C.H. Joiner, and P.A. Lane, Hydroxyurea effectiveness in children and adolescents with sickle cell anemia: A large retrospective, population-based cohort. Am J Hematol, 2017. 92(1): p. 77–81.

14. Tshilolo, L., G. Tomlinson, T.N. Williams, B. Santos, P. Olupot-Olupot, A. Lane, B. Aygun, S.E. Stuber, T.S. Latham, P.T. McGann, and R.E. Ware, Hydroxyurea for Children with Sickle Cell Anemia in Sub-Saharan Africa. New England Journal of Medicine, 2018. 380(2): p. 121–131.

15. Tubman, V.N., R. Marshall, W. Jallah, D. Guo, C. Ma, K. Ohene-Frempong, W.B. London, and M.M. Heeney, Newborn Screening for Sickle Cell Disease in Liberia: A Pilot Study. Pediatr Blood Cancer, 2016. 63(4): p. 671–6.

16. Williams, S.A., B. Browne-Ferdinand, Y. Smart, K. Morella, S.G. Reed, and J. Kanter, Newborn Screening for Sickle Cell Disease in St. Vincent and the Grenadines: Results of a Pilot Newborn Screening Program. Glob Pediatr Health, 2017. 4: p. 2333794x17739191.

17. Therrell, B.L., Jr., M.A. Lloyd-Puryear, J.R. Eckman, and M.Y. Mann, Newborn screening for sickle cell diseases in the United States: A review of data spanning 2 decades. Semin Perinatol, 2015. 39(3): p. 238–51.

18. Aygun, B. and I. Odame, A global perspective on sickle cell disease. Pediatric Blood & Cancer, 2012. 59(2): p. 386–390.

19. Allaf, B., F. Patin, J. Elion, and N. Couque, New approach to accurate interpretation of sickle cell disease newborn screening by applying multiple of median cutoffs and ratios. Pediatr Blood Cancer, 2018. 65(9): p. e27230.

20. Nagel, R.L., M.E. Fabry, and M.H. Steinberg, The paradox of hemoglobin SC disease. Blood Rev, 2003. 17(3): p. 167–78.

21. An, R., M.N. Hasan, Y. Man, and U.A. Gurkan, Integrated Anemia Detection and Hemoglobin Variant Identification Using Point-of-Care Microchip Electrophoresis. Blood, 2019. 134(Supplement_1): p. 378-378.

22. Fraiwan, A., M.N. Hasan, R. An, J.Z. Xu, A.J. Rezac, N.J. Kocmich, T. Oginni, G.M. Olanipekun, F. Hassan-Hanga, B.W. Jibir, S. Gambo, A.K. Verma, P.K. Bharti, S. Riolueang, T. Ngimhung, T. Suksangpleng, P. Thota, R. Shanmugam, A. Das, V. Viprakasit, C.M. Piccone, J.A. Little, S.K. Obaro, and U.A. Gurkan, International Multi-Site Clinical Validation of Point-of-Care Microchip Electrophoresis Test for Hemoglobin Variant Identification. Blood, 2019. 134(Supplement_1): p. 3373-3373.

23. Canning, D.M. and R.G. Huntsman, An assessment of Sickledex as an alternative to the sickling test. Journal of clinical pathology, 1970. 23(8): p. 736–737.

24. Steele, C., A. Sinski, J. Asibey, M.D. Hardy-Dessources, G. Elana, C. Brennan, I. Odame, C. Hoppe, M. Geisberg, E. Serrao, and C.T. Quinn, Point-of-care screening for sickle cell disease in low-resource settings: A multi-center evaluation of HemoTypeSC, a novel rapid test. Am J Hematol, 2019. 94(1): p. 39–45.

25. Kanter, J., M.J. Telen, C. Hoppe, C.L. Roberts, J.S. Kim, and X. Yang, Validation of a novel point of care testing device for sickle cell disease. BMC Medicine, 2015. 13(1): p. 225.

26. Eastman, J.W., R. Wong, C.L. Liao, and D.R. Morales, Automated HPLC screening of newborns for sickle cell anemia and other hemoglobinopathies. Clinical Chemistry, 1996. 42(5): p. 704–710.

27. Ross, L.F., Mandatory versus voluntary consent for newborn screening? Kennedy Inst Ethics J, 2010. 20(4): p. 299–328.

28. Organization, G.W.H., First WHO Model List of Essential In Vitro Diagnostics. 2019 ((WHO Technical Report Series, No. 1017)).

29. Shinkins, B., M. Thompson, S. Mallett, and R. Perera, Diagnostic accuracy studies: how to report and analyse inconclusive test results. BMJ: British Medical Journal, 2013. 346: p. f2778.

30. Bossuyt, P.M., J.B. Reitsma, D.E. Bruns, C.A. Gatsonis, P.P. Glasziou, L. Irwig, J.G. Lijmer, D. Moher, D. Rennie, H.C.W. de Vet, H.Y. Kressel, N. Rifai, R.M. Golub, D.G. Altman, L. Hooft, D.A. Korevaar, J.F. Cohen, and f.t.S. Group, STARD 2015: An Updated List of Essential Items for Reporting Diagnostic Accuracy Studies. Clinical Chemistry, 2015. 61(12): p. 1446–1452.

31. Kutlar, A., F. Kutlar, J.B. Wilson, M.G. Headlee, and T.H.J. Huisman, Quantitation of hemoglobin components by high-performance cation-exchange liquid chromatography: Its use in diagnosis and in the assessment of cellular distribution of hemoglobin variants. American Journal of Hematology, 1984. 17(1): p. 39–53.

32. Thomas, C. and A.B. Lumb, Physiology of haemoglobin. Continuing Education in Anaesthesia Critical Care & Pain, 2012. 12(5): p. 251–256.

33. Metaxotou-Mavromati, A.D., H.K. Antonopoulou, S.S. Laskari, H.K. Tsiarta, V.A. Ladis, and C.A. Kattamis, Developmental Changes in Hemoglobin F Levels During the First Two Years of Life in Normal and Heterozygous β-Thalassemia Infants. Pediatrics, 1982. 69(6): p. 734–738.

34. Ryan, K., B.J. Bain, D. Worthington, J. James, D. Plews, A. Mason, D. Roper, D.C. Rees, B.d.l. Salle, and A. Streetly, Significant haemoglobinopathies: guidelines for screening and diagnosis. British Journal of Haematology, 2010. 149(1): p. 35–49.

35. Keren, D.F., D. Hedstrom, R. Gulbranson, C.-N. Ou, and R. Bak, Comparison of Sebia Capillarys Capillary Electrophoresis With the Primus High-Pressure Liquid Chromatography in the Evaluation of Hemoglobinopathies. American Journal of Clinical Pathology, 2008. 130(5): p. 824–831.

36. Borbely, N., L. Phelan, R. Szydlo, and B. Bain, Capillary zone electrophoresis for haemoglobinopathy diagnosis. Journal of Clinical Pathology, 2013. 66(1): p. 29–39.

37. Nusrat, M., B. Moiz, A. Nasir, and M. Rasool Hashmi, An insight into the suspected HbA2’ cases detected by high performance liquid chromatography in Pakistan. BMC Research Notes, 2011. 4(1): p. 103.

38. Sharma, P. and R. Das, Cation-exchange high-performance liquid chromatography for variant hemoglobins and HbF/A2: What must hematopathologists know about methodology? World journal of methodology, 2016. 6(1): p. 20–24.

39. Kreuels, B., C. Kreuzberg, R. Kobbe, M. Ayim-Akonor, P. Apiah-Thompson, B. Thompson, C. Ehmen, S. Adjei, I. Langefeld, O. Adjei, and J. May, Differing effects of HbS and HbC traits on uncomplicated falciparum malaria, anemia, and child growth. Blood, 2010. 115(22): p. 4551–4558.

40. Fucharoen, S. and D.J. Weatherall The hemoglobin E thalassemias. Cold Spring Harbor perspectives in medicine, 2012. 2, DOI: 10.1101/cshperspect.a011734.

41. Warghade, S., J. Britto, R. Haryan, T. Dalvi, R. Bendre, P. Chheda, S. Matkar, Y. Salunkhe, M. Chanekar, and N. Shah, Prevalence of hemoglobin variants and hemoglobinopathies using cation-exchange high-performance liquid chromatography in central reference laboratory of India: A report of 65779 cases. Journal of laboratory physicians, 2018. 10(1): p. 73–79.

42. Masiello, D., M.M. Heeney, A.H. Adewoye, S.H. Eung, H.-Y. Luo, M.H. Steinberg, and D.H.K. Chui, Hemoglobin SE disease—A concise review. American Journal of Hematology, 2007. 82(7): p. 643–649.

43. Xu, J.Z., S. Riolueang, W. Glomglao, K. Tachavanich, T. Suksangpleng, S. Ekwattanakit, and V. Viprakasit, The origin of sickle cell disease in Thailand. International Journal of Laboratory Hematology, 2019. 41(1): p. e13–e16.

44. Colah, R., A. Gorakshakar, and A. Nadkarni, Global burden, distribution and prevention of β-thalassemias and hemoglobin E disorders. Expert Review of Hematology, 2010. 3(1): p. 103–117.

